# Granulocyte-macrophage colony-stimulating factor (GM-CSF) antibodies treatment for COVID-19 patients: a meta-analysis

**DOI:** 10.1101/2022.01.07.22268878

**Authors:** Jin-Tao Guan, Wei-Jie Wang, Anran Xi, Du Jin, Xiao-Yue Mou, Zheng-Hao Xu

## Abstract

**Objective:** We performed a meta-analysis in order to determine safety of granulocyte-macrophage colony-stimulating factor (GM-CSF) antibodies on COVID-19.

**Methods:** We searched from the Cochrane Library, PubMed, Embase, biorxiv and medrxiv databases beginning in the COVID-19 outbreak on December 1, 2019 until August 29, 2021. The primary outcomes included death, the incidence of invasive mechanical ventilation (IMV), ventilation requirement, and secondary infection.

**Results:** 6 eligible literature involving 1501 COVID-19 patients were recruited, and they were divided into experimental group (n = 736) and control group (n = 765). Using a random-effect model, we found that the GM-CSF antibodies treatment was associated with a 3.8-26.9% decline of the risk of mortality[odd’s ratio (OR) = 0.06, 95% confidence interval (CI): -0.11, -0.01, p =0.02], a 5.3-28.7% reduction of incidence of IMV [OR = 0.51, 95% CI: 0.28, 0.95, p =0.03], and a 23.3-50.0% enhancement of ventilation improvement [OR = 11.70, 95% CI: 1.99, 68.68, p=0.006]. There were no statistically significant differences in the association between two groups in second infection.

**Conclusion:** Severe COVID-19 patients may benefit from GM-CSF antibodies.

## Introduction

The COVID-19 pandemic is still threatening public health by its serious outcomes and strong infectivity The classic symptoms of COVID-19 include high fever, respiratory failure, and even acute respiratory distress syndrome (ARDS). Granulocyte-macrophage colony-stimulating factor (GM-CSF) antibodies inhibiting GM-CSF signaling axis by targeting GM-CSF, such as Gimsilumab, lenzilumab, namilumab, and otilimab, or GM-CSF receptors (GM-CSFR), such as Mavrilimumab. Though none of anti-GM-CSF or anti-GM-CSFR antibody are currently FDA-approved for regular medical use, GM-CSFR antibodies may be beneficial to autoimmune and inflammatory disorders in patients, such as rheumatoid arthritis. GM-CSF plays an important role in the pathogenesis of COVID-19 for its immune hyper response. Therefore, anti-GM-CSF therapy has been applied to hospitalized patients with severe COVID-19 and has achieved certain results^1-3^. In the announced clinical trials, GM-CSF antibodies, including mavrilimumab [ClinicalTrials.gov identifier: NCT04492514], lenzilumab [ClinicalTrials.gov identifier: NCT04351152] have been used in the treatment of COVID-19. However, their underlying pharmacological mechanisms, safety, and adverse events in the treatment of COVID-19 have not been clearly clarified. In addition, the safety and efficacy of GM-CSF antibodies in the treatment of COVID-19 are also controversial; in particular, mortality, drug use, and secondary infections should be of concern^4–6^. Therefore, the present meta-analysis was conducted to investigate GM-CSF antibody treatment in COVID-19 patients.

## Methods

This study conducted according to the Preferred Reporting Items for Systematic reviews and Meta-Analyses (PRISMA) and registered in PROSPERO (CRD42020221450; https://www.crd.york.ac.uk/prospero/).

### Inclusion and Exclusion Criteria and Data Collection

The literature search and data analyses were undertaked by J.G. and W.W, Then, the duplicate literatures were excluded using EndNote X7; Only eligible literature with full text included. Any disagreement was resolved by the third investigator (A.X). Briefly, literature concerning GM-CSF antibody treatment alone, or in combination with other specific treatments, of adult COVID-19 patients were included. Simultaneously, the studies were Research in children and which were without clear results were excluded.

### Search strategy

PRISMA guidelines were based on the PRISMA 2020 checklist.. The search strategy was ran by J.G. and W.W. in the Cochrane Library, Embase, PubMed, and biorxiv and medrxiv. References for each piece of literature were manually reviewed. Any disagreement was solved by the third author (Z.X.). In detail, relevant literature published from 1 December 2019 and until 29 August 2021 with the following keywords were searched: (“SARS-CoV-2” OR “coronavirus” OR “nCoV” OR “pneumonia” OR “corona-virus” OR “2019nCoV” OR “COVID-19”) AND (“lenzilumab” OR “TJM” OR “recombinant monoclonal antibodies against granulocyte macrophage colony-stimulating factor” OR “monoclonal antibodies against granulocyte macrophage colony-stimulating factor” OR “antibodies against granulocyte macrophage colony-stimulating factor” OR “mavrilimumab” OR “Otilimab” OR “TJ003234” OR “Roivant” OR “MOR103” OR “kb003” OR “plonmarlimab” OR “granulocyte macrophage colony stimulating factor” OR “GM-CSF” OR “CSF”). Of note, preprint results from registered clinical trials were also included.

### Study selection and Data extraction

J. G. and W.W were responsible for extracting data through eligible literature, including the number of recruited patients, therapeutic strategies, ventilation conditions, ICU admission risk, death number, severe case number, risks of COVID-19, length of stay, secondary infection, and severe events (e.g., sepsis, acute kidney injury, cardiac injury), etc.

### Assessment of study quality

Study quality was assessed using the Newcastle-Ottawa Scale (NOS) ^7^ (**Supplemental Table 1**) for nonrandomized studies and the Cochrane risk-of-bias tool (**Supplemental Figure 1**) for randomized control trials (RCTs).

### Statistical analysis

Pooled estimates were presented as odds ratios (OR) and 95% confidence intervals (CIs) and visualized by forest plots. As the studies included in the analysis are not functionally identical (such as the use of different kinds of drugs and different standard of care methods), a random-effects model (Mantel–Haenszel method) was employed for the meta-analysis. Heterogeneity among studies was evaluated by χ^2^, I^2^, df, and Tau^2^. Negative I^2^ values were set to zero. 25%, 50%, and 75% I^2^ indicated a low, moderate, and high heterogeneity, respectively. Publication bias was assessed by using funnel plots and Egger’s asymmetry test. Revman 5.3 was used for statistical processing. In addition, to evaluate the strength and stability of the meta-analysis, sensitivity analysis was conducted by omitting the individual studies one by one.

## Result

### Selection of eligible literatures

Following the searching strategy described in Figure 1, 1967 pieces of literature were initially identified based on the assessment of the titles and abstracts. We excluded 1961 pieces of literature strictly conformed to the inclusion and exclusion criteria. At last, a total of 6 pieces of eligible literature were finally included in the meta-analysis.

**Figure 1.**
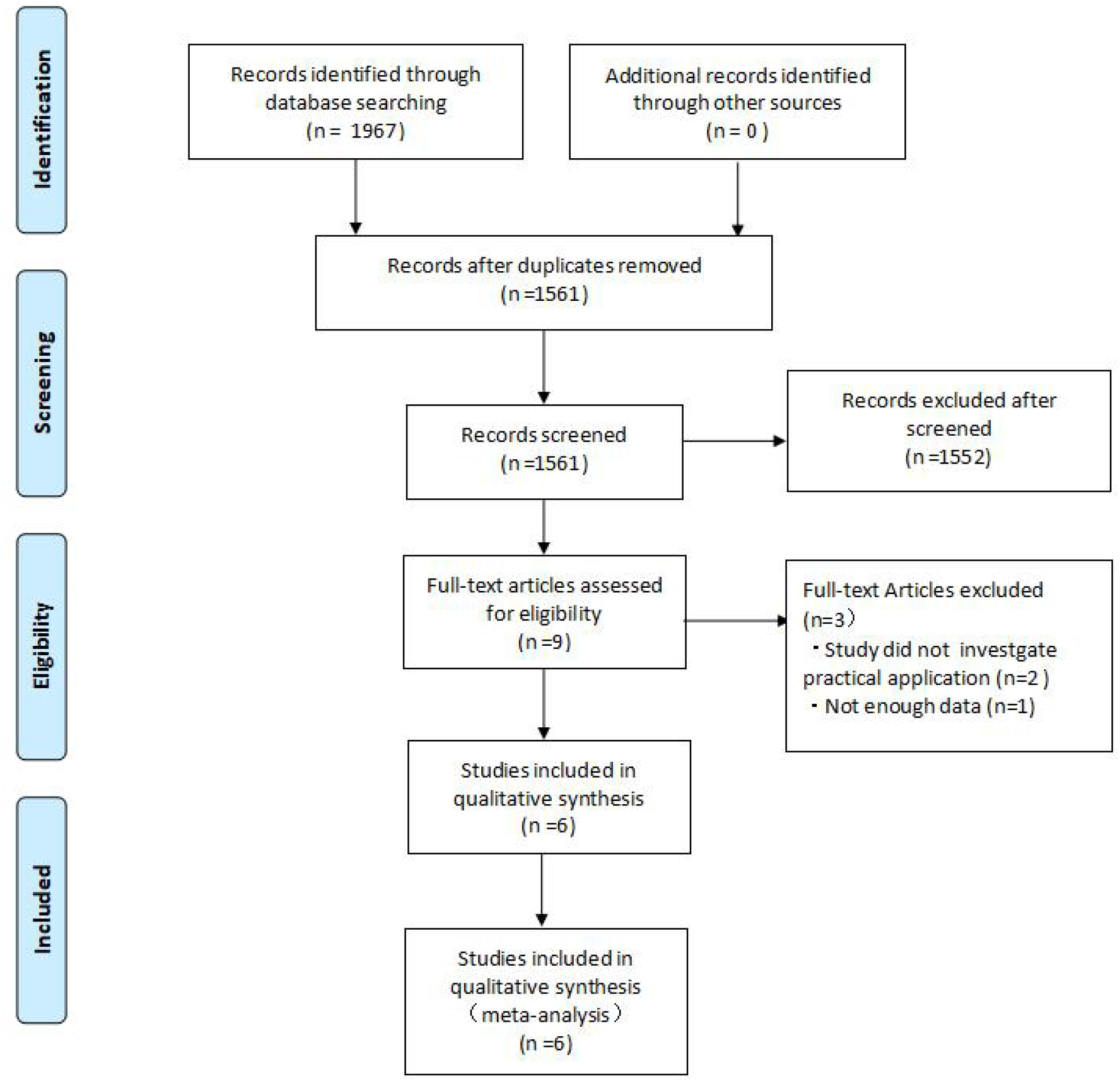
Flow diagram of selection of eligible literatures.

### Characteristics of included literatures

A total of 6 pieces of eligible literature^8-13^ involving 1501 COVID-19 patients were divided into the GM-CSF antibody treatment group (n = 736) and control group (n = 765). Two of included studies were nonrandomized studies and the other four were RCTs. Characteristics of the included studies were summarized in **Table 1**, including the study design (type of study), population, treatment methods, sample sizes, age, gender, standard of care, primary endpoints, and Secondary endpoints. The NOS scores of included non-RCTs were shown in Supplementary Table 1 and risk of bias of included RCTs were shown in Supplementary Figure 1.

### Mortality

Mortality data were obtained from 6 studies involving 1501 COVID-19 patients; of these, 736/1501received GM-CSF antibody treatment. As shown in Figure 2, GM-CSF antibody treatment significantly improved the overall survival (OS) of COVID-19 than conventional treatment (OR = 0.06, 95% CI: -0.11, -0.01). The test for the overall efficacy manifested as the following: Z = 2.38 (p= 0.02), Tau ^2^ = 0.00, Chi ^2^ = 6.70, df =5 (p = 0.24), and I^2^ = 25%). In subgroup analysis, the finding from two small cohort studies (non-RCTs) indicated GM-CSF antibody therapy did not affect mortality (OR = -0.19; 95% CI: -0.36,-0.03); However, the finding based on four RCTs indicated that GM-CSF antibody treatment reduce the mortality (OR = -0.06; 95% CI: -0.11,-0.01).

**Figure 2.**
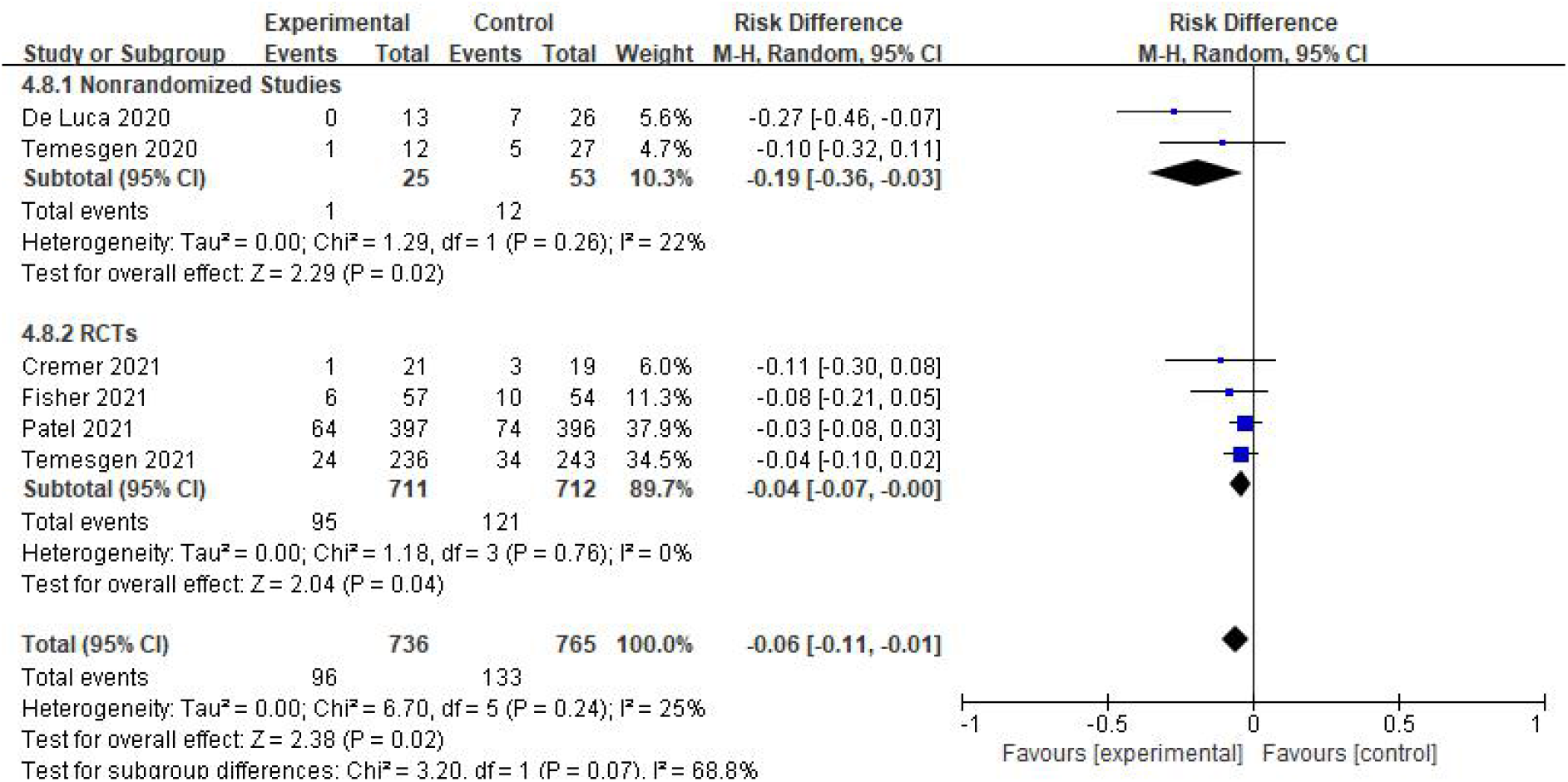
Risk of mortality between GM-CSF antibodies and control groups

### Incidence of invasive mechanical ventilation (IMV)

To estimate the risk of Incidence of IMV in COVID-19 atients, we analyzed the data of 597 COVID-19 patients from 4 studies, including 282 in the GM-CSF antibody treatment group and 315 in the control group. In the former group, 37/282 patients were admitted for incidence of IMV; while 71/315 in the control group were admitted for incidence of IMV. As shown in Figure 3, GM-CSF antibody treatment significantly reduce incidence of IMV (OR = 0.51; 95% CI: 0.28, 0.95). The test for the overall efficacy was Z =2.12 (p = 0.03), Tau^2^ = 0.06, Chi^2^ = 3.24, df = 3 (p = 0.36), and I^2^ = 7%. This finding was also replicated by subgroup analysis of both cohort studies (OR = 0.16; 95% CI: 0.03, 0.74) and RCTs (OR = 0.64; 95% CI: 0.40, 1.03).

**Figure 3.**
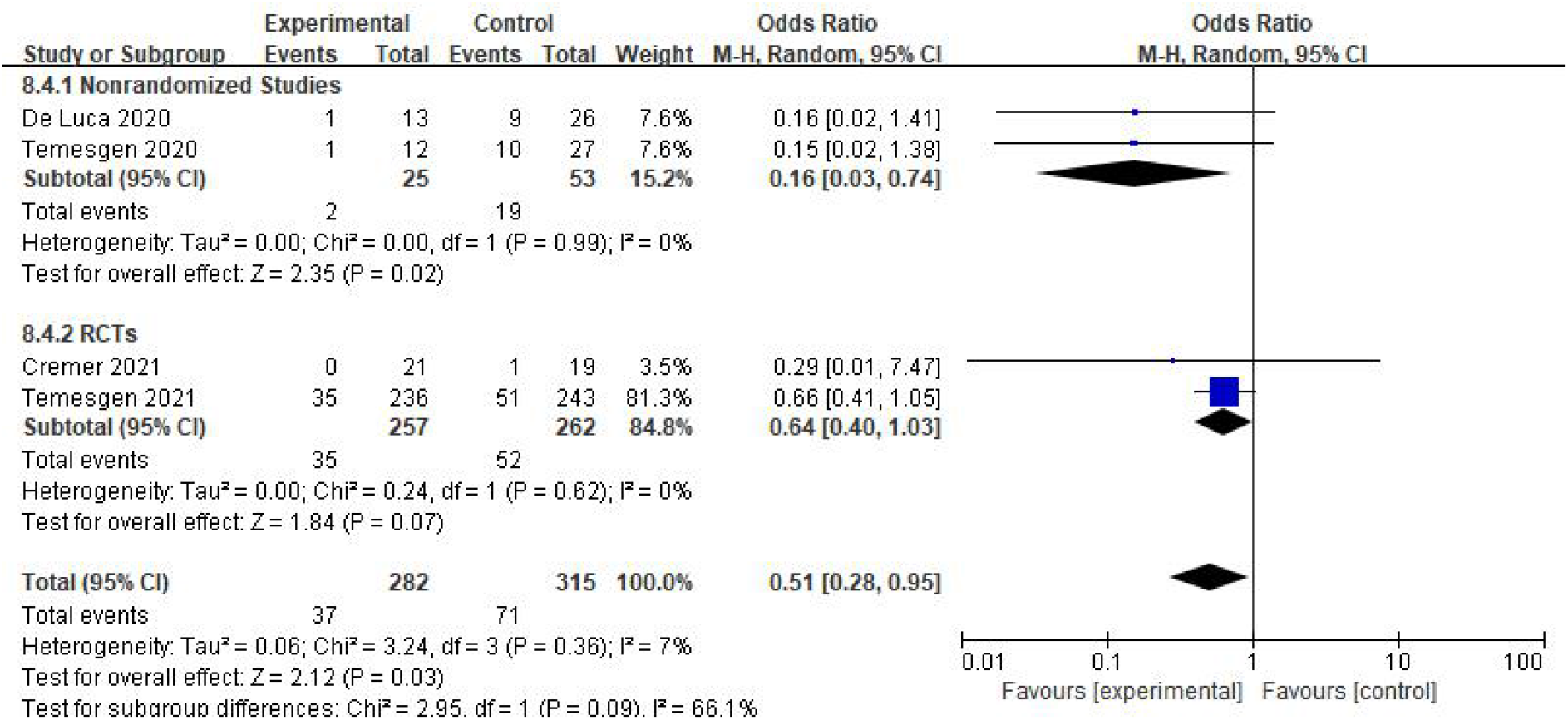
The risk of IMV between the GM-CSF antibody treatment group and the control group in COVID-19 patients.

### Improvement of ventilation

Ventilation data (need oxygen therapy to without oxygen therapy) from 2 controlled clinical trials involving 79 COVID-19 patients (34 in the GM-CSF antibody treatment group and 45 in the control group) were analyzed to assess the overall improvement on ventilation. A total of 19/34 in the GM-CSF antibody treatment group and 14/45 patients in control group had improved ventilation, respectively. As shown in Figure 4, the treatment of GM-CSF antibodies improved the ventilation in COVID-19 patients (OR = 11.70, 95% CI: 1.99, 68.68). The test for the overall efficacy was Z = 2.72 (p = 0.006), Tau^2^ = 0.00, Chi ^2^ = 0.52, df = 1 (p = 0.47), and I ^2^ = 0%. This finding was also replicated by subgroup analysis of both cohort studies (OR = 27.0; 95% CI: 1.45, 501.49) but not RCTs (OR = 7.20; 95% CI: 0.78, 66.63).

**Figure 4.**
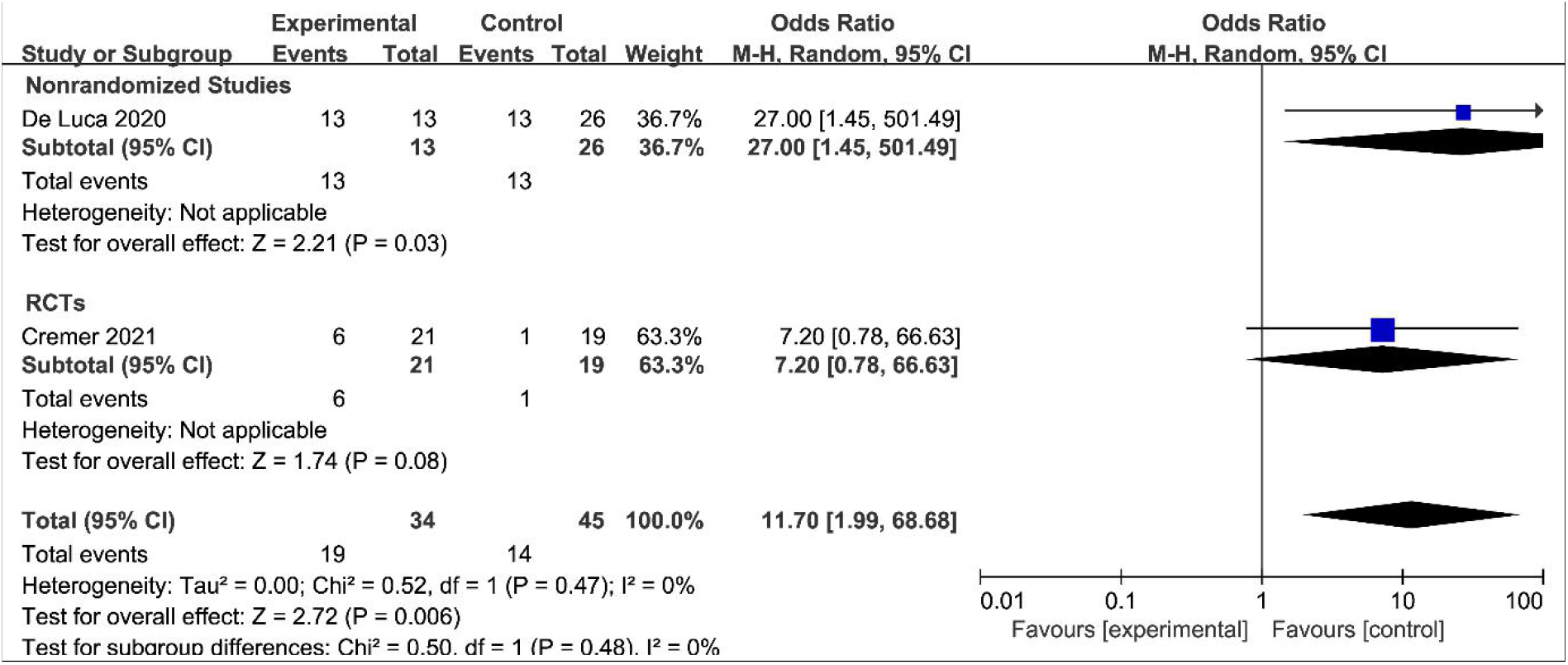
Improvement of ventilation between GM-CSF antibodies and control groups COVID-19 patients

### Secondary infection

Secondary infections were reported in 5 studies involving 1462 COVID-19 patients; of these, 724 were in the GM-CSF antibody treatment group and 738 were in the control group. As shown in Figure 5, The data did not reveal a significant difference in secondary infection between groups (OR = 0.82, 95% CI: 0.58, 1.15). The test for the overall efficacy was Z = 1.15 (p = 0.25), Tau^2^ = 0.00, Chi^2^ = 1.73, df = 4 (p = 0.78), and I^2^ = 0%.

**Figure 5.**
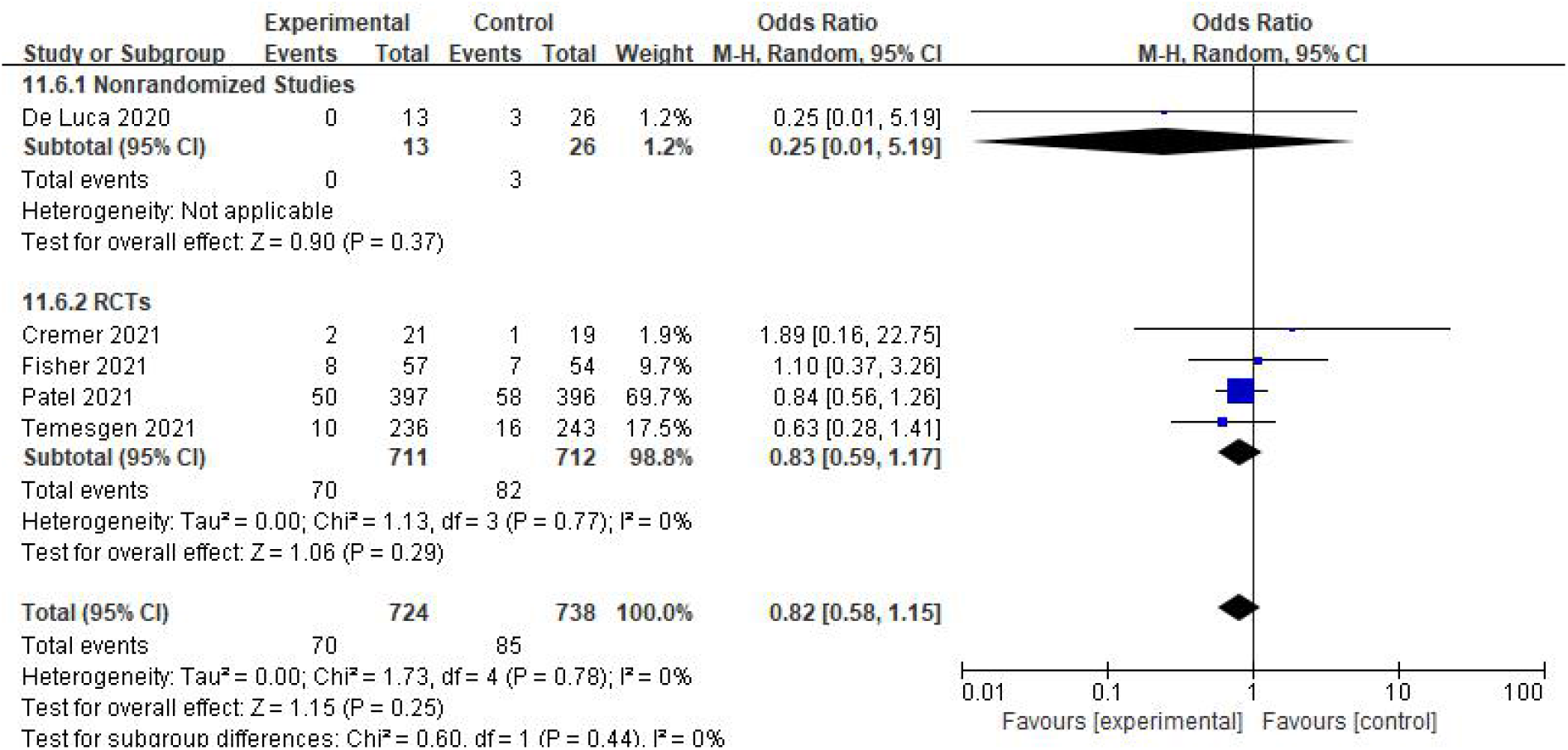
The risk of secondary infection between the GM-CSF antibody treatment group and the control group in COVID-19 patients.

## Discussion

The COVID-19 pandemic is still an urgent problem in the world and the cases continue to rise globally. Recent studies revealed that GM-CSF can enhance the expression of many pro-inflammatory cytokines and chemokines, such as IL-1, IL-6, and TNF.^14^ The anti-GM-CSF monoclonal antibody has been approved emergency compassionate use for patients with COVID-19 by the US Food and Drug Administration.^15^ The anti-GM-CSF monoclonal antibody might have broader effects than other immunomodulatory approaches on the systemic pro-inflammatory responses accompanying the cytokine release syndrome in COVID-19.

Based on the considerable potential of GM-CSF antibodies to treat COVID-19, this article intends to evaluate the clinical efficacy and safety of GM-CSFantibodies for COVID-19. Our meta-analysis involving 6 studies about GM-CSF antibodies treat in COVID-19 patients. Categorized by the therapeutic strategies, they were divided into the GM-CSF antibody treatment group (n = 736) and the control group (n = 765). Our results revealed that GM-CSF antibodies treatment reduced mortality, decreased the incidence of IMV, and improve the ventilation in COVID-19 patients. Of note, Patel et al.^8^ found otilimab has a substantial benefit in patients aged ≥ 70, possibly reflecting a population that could benefit from therapeutic blocking of GM-CSF in severe COVID-19 patients.. Taken together, some other factors, such as the age, may still influence the efficacy of GM-CSF antibodies treatment, though our data provide a relatively high quality evidence to support that GM-CSF antibody treatment for reducing Mortality in COVID-19 patients.

The lastly case repored that GM-CSF antibodies may be beneficial to patients who needed supplemental oxygen, and failed multiple therapy. ^16^, which may be related to the decreased ventilation risk. COVID-19 progressively causes severe respiratory failure and serious inflammation, as a result, this leads to high mortality risk. However, Rodríguez-Molinero et al.^17^ believed that early intervention of anti-inflammatory (interleukin 6 inhibition) also triggers adverse events in COVID-19 patients. Kimmig et al.^18^ and Quartuccio et al.^19^ revealed that COVID-19 patients in the anti-inflammatory treatment (interleukin 6 inhibition) group have a higher rate of secondary infection. In GM-CSF inhibition study, we did not obtain a significant difference in secondary infection in COVID-19 patients between GM-CSF antibodies treatment group and control, which may be attributed that they already have a high incidence of secondary infections.^20^ Consistent with previous meta-analyses, unmeasured confounding factors and potential biases in our study should be a concern. Therefore, our primary analysis provided consistent results across most analyses. Although adjustments for potential confounders were performed in our study, some unmeasured confounding factors may exist. In addition, our study may include missing data for some variables and there is potential for inaccuracies in the documentation of electronic health records. Taken together, this meta-analysis provided a systematic comprehensive, and updated evaluation of GM-CSF antibodies treatment in COVID-19-related clinical outcomes.

Several limitations in our meta-analysis should be noted as follows: (1) because of limited number of cases and studies, the quality of all cohort-study-based evidences and most RTC-based evidences are low, especially for the improvement of ventilation and the incidence of IMV; We believe that low-powered analysis of this parameter based on a small number of studies can still provide useful insights by highlighting a deficiency in a particular topic that deserves further attention. (2) There are differences in the standard of care used (for placebo/GM-CSF inhibitor arms) in the included studies, which limits our capability to perform an adequate comparison and meta-analysis of these studies. For example, differences in the oxygen therapy and nursing measures may influence the mortality in COVID-19 patients. (3) One of the six studies used for the meta-analysis are pre-publications without peer-review, which may imply a risk of publication bias or other potential risks. (4) Study population is different for some studies, which implies a risk of selection bias; (5) Besides, there are some other factors in this study, such as other drugs, ventilator availability, and vaccines. For example, the effectiveness of dexamethasone in COVID-19 has been previously reported.^21^ So, the synergistic effect or other interactions of these factors (such as dexamethasone) remains unclear. Therefore, the clinical interpretation of these findings is limited by these high or unclear risks of bias. More RCTs and cohort studies are still needed for further verification. Especially, further large and multi-center clinical studies are still warranted.

## Conclusions

Due to the urgent demand for effective treatments for COVID-19, this meta-analysis study comprehensively analyzed the safety and efficacy of GM-CSF antibody treatment, which was identified as being beneficial to COVID-19 patients. Different types of GM-CSF antibodies administration are currently being therapeutically tested in COVID-19 clinical trials. The application of GM-CSF anti-bodies can reduce respiratory symptoms; however, evidence supporting the function of GM-CSF anti-bodies in reducing secondary infections of COVID-19 is limited. Besides, our study had some limitations since two observational studies were included; potential biases and confounding factors could not be excluded. Therefore, more RCTs and high-quality literature is required to validate our study. Especially, several medications in the past have been highlighted and even received emergency approvals based on flawed studies in the age of COVID-19, which may ultimately lead to drug shortages for patients who really need those medications.

## Data Availability

All data produced in the present work are contained in the manuscript

## Author contributions

The study was designed by J.G. J.G and W.W. ran the search strategies and undertook the search.. J.D and M.X extracted data and analysis. J.G. and Z.X. wrote the manuscript. J.G., and W.W. did the correction under the supervision of Z.X. A.X. and J.G. contribute to the revision for collected data, edited the manuscript, and check all the extracted data under the supervision of Z.X.

## Consent statement and ethical approval

Consent statement and ethical approval are not required as the current study was based on published data.

## Availability of data, code and other materials

All data are presented in the text.

